# New Genetic Insights in Rheumatoid Arthritis using Taxonomy3^®^, a Novel method for Analysing Human Genetic Data

**DOI:** 10.1101/2023.02.21.23286176

**Authors:** Justyna Kozlowska, Neil Humphryes-Kirilov, Anastasia Pavlovets, Martin Connolly, Zhana Kuncheva, Jonathan Horner, Ana Sousa Manso, Clare Murray, J. Craig Fox, Alun McCarthy

## Abstract

Genetic support for a drug target has been shown to increase the probability of success in drug development, with the potential to reduce attrition in the pharmaceutical industry alongside discovering novel therapeutic targets. It is therefore important to maximise the detection of genetic associations that affect disease susceptibility. Conventional statistical methods used to analyse genome-wide association studies (GWAS) only identify some of the genetic contribution to disease, so novel analytical approaches are required to extract additional insights. C4X Discovery has developed a new method Taxonomy3^®^ for analysing genetic datasets based on novel mathematics. When applied to a previously published rheumatoid arthritis GWAS dataset, Taxonomy3^®^ identified many additional novel genetic signals associated with this autoimmune disease. Follow-up studies using tool compounds support the utility of the method in identifying novel biology and tractable drug targets with genetic support for further investigation.

## Introduction

Attrition in drug development is a major issue for the pharmaceutical industry, particularly for complex diseases, with significant R&D resources spent on projects that do not deliver new medicines for patients. It is now widely accepted that genetic support of a therapeutic drug target leads to an increased probability of successful delivery to patients of a new medicine, with the potential to significantly reduce attrition ^1,2^. These observations have directed the focus of drug discovery groups on results from genetic studies of disease. As opposed to rare diseases, which are often caused by the dysfunction of a single gene, common diseases are complex traits influenced by the added contribution of many genetic variants. Conventional analysis of Genome-wide association studies (GWAS) have generated thousands of associations, initially based on the study of single-nucleotide polymorphisms (SNPs) arrays but increasingly using whole exome - or whole genome-sequence data ^3^. A critically important observation is that >90% of GWAS variants identified fall in non-coding regions of the genome and thus do not directly affect the coding sequence of a gene but rather accumulate in DNA regulatory elements and can disrupt binding sites for transcription factors likely regulating the expression levels of genes in a cell type-specific manner. Therefore, it is unclear which genes these variants regulate and in which cell types or physiological contexts this regulation occurs. This has hindered the translation of GWAS findings and insights into clinical interventions ^4^.

Another key current limitation of using GWAS datasets to provide insights on relevant disease biology is that no analysis method can extract all the genetic information relevant to a disease embedded in the genetics, leading to the concept of ‘missing heritability’ i.e. the genetic component of disease susceptibility that is calculated from family or twin studies, but cannot be fully accounted for by the known genetic associations detected.

Many explanations for this missing heritability have been proposed, including: the existence of many rare mutations with strong effect not captured by the standard GWAS analysis methodologies ^5^ ; gene-gene interactions with strong effect (where the individual genes are not in themselves sufficiently strongly associated with the disease to be detected) ^6–8^; or ‘omnigenic’ models proposing that many thousands of genes with small effect impact disease susceptibility such that even current meta-analyses are insufficiently powered to detect most of the signals ^6–8^.

Based on twin and family studies, heritability of rheumatoid arthritis (RA) is estimated at ∼60%, showing a significant impact of genetic variation on disease aetiology ^9^. A number of RA GWAS have been published, with datasets that have included over 300,000 people ^10–12^. The largest of these studies identified ∼75 loci associated with diagnosis of RA (11 of which were novel) and while this is an important contribution to understanding the aetiology of the disease, the authors calculate that the total of all these findings accounts for 40-50% of the heritability thought to affect development of RA, leaving significant missing heritability still to be identified ^12^.

Whatever the reasons for the missing heritability, it is clear that many different analytical approaches are required to maximise the extraction of genetic insights from disease datasets. To this end, C4X Discovery has developed a unique method (called Taxonomy3^®^) for analysing human genetic datasets based on applying novel mathematics. The core of the method converts genotypes into numbers that retain the information content of the genotypes in the context of the case/control distinction being studied. This conversion allows linear algebra methods to be applied directly to the data, probing the data in new ways. In this paper, we present details of application of Taxonomy3^®^ to a rheumatoid arthritis case/control dataset, show the additional genetic insights generated by the method, and provide examples where the findings allow the potential initiation of drug discovery programmes based on drug targets identified through these new genetic findings.

## Methods

### Overview of Taxonomy3^®^ Method

Taxonomy3^®^ is based on correlations of individualized divergences (named Log Bayes Factors, LBFs) ^13^, and their Eigen decomposition: the mathematical principle of the method has been described previously ^14–16^. Briefly, for each genotype in a case/control group, the LBF is calculated, and for all subjects, each genotype is replaced by the calculated LBF for that genotype. In this way, the initial genotype data matrix is transformed into a LBF matrix of the same dimension, representing the information gain provided by each subject and SNP pertaining to the overall case/control distinction. LBFs have additive properties allowing the use of linear algebra tools. Eigen decomposition of correlations of LBFs (PCA) is the preferred multivariate analysis method as it produces independent sets of correlated variables. To accurately determine how much each variable contributes to case/control distinction, we projected variable loadings on to the observed case/control direction in relevant dimensional space. Other datatypes (gene expression, clinical variables etc.) can also be transformed into LBFs, which in turn can be co-analysed alongside genotype LBFs ^15^.

The principal component results are visualized using biplots to display the relative position of subjects and variables. From this, variables can be identified that are important for discriminating cases from controls. Statistical significance is assessed by permutation of the case/control labels and re-analysis. For each variable, p-values are obtained by comparing the true observer projected loading to the Gaussian mixture model (Mixmod software^17^) fitted to the distribution of the permuted projected loadings. The Family Wise Error Rate Šidák correction for multiple testing is used to define the genome-wide threshold for significant variables using the exact number of tests being carried out.

All analyses were performed with proprietary software on a cluster of Linux machines (Amazon Web Services). The software has had various iterations and the latest production code has been developed in C++ and CUDA and passes an exhaustive set of tests.

### Taxonomy3^®^ Analysis Data Management

Before running the Taxonomy3^®^ analysis of the case/control datasets, subjects and variables with poor quality data were excluded, and Taxonomy3^®^ co-analysis with HapMap populations used to identify case and control cohorts for analysis that were matched for genetic ancestry.

### Subjects and Phenotype Data

We analysed the rheumatoid arthritis (RA) dataset from the Welcome Trust Case Control Consortium (WTCCC), comprising RA cases (n=1999) recruited from sites across the UK ^11^. As controls, we studied UK National Blood Service (NBS) controls (n=1480) from the same source. Both cases and controls were genotyped with the Affymetrix 500K SNP chip (For more information on the cases and controls, see ^11^. The WTCCC has limited phenotype data on the disease samples: disease status, age, sex and broad geographical region within Britain. The downloaded data were stored and analysed on the Amazon Web Services cloud computing facilities. To ensure security, the data were encrypted, stored behind a HIPAA-compliant firewall, and access to the data was restricted to a defined list of IP addresses.

### Genotypes

The latest annotations file for the discontinued Affymetrix 500K DNA chip was obtained from the manufacturer. We used genotypes derived using the Chiamo algorithm as in the original WTCCC analysis, discarding genotypes having a call probability lower than 90% ^11^.

### Univariate Quality Control

The case and control data were first subject to conventional data quality control. The objective of this procedure was to detect and remove potential biases that could undermine the analysis.

Gender: Checking for gender discrepancies is critical to Taxonomy3^®^ as LBFs are not defined in the same way for X-linked SNPs in males and females. Gender was inferred for each patient from intensities of X-linked genotypes and X chromosome heterozygosity, and checked against the reported gender.

Missing values: Subjects with a missing value rate above 5% were removed from the analysis.

Relatedness: The objective of this analysis was to detect and remove inter-related subjects/samples from subsequent analyses. We selected a total of 35,941 highly variable SNPs having a high Minor Allele Frequency (above 48%). We then determined the percentage of identical genotypes in all possible pairs of subjects.

Variables: Variables were excluded from the analysis for any of the following reasons:

1. Monomorphic in the whole population
2. Missing >5% of values
3. SNPs departing from Hardy Weinberg Equilibrium in the control group (threshold p=10^−8^). In classical GWAS, SNPs having a low minor allele frequency (MAF) are usually removed due to power considerations. This is not necessary in Taxonomy3^®^, as the method can handle rare variants.

### HLA Imputation

Human leukocyte (HLA) genotyping is not available in WTCCC RA dataset, therefore imputation of antigen (HLA) genotypes from SNP genotype data was performed in-house using the HIBAG imputation method ^18^ with pre-trained parameter estimates specific to Affymetrix500k for European ancestry.

### Multivariate Quality Control

As Taxonomy3^®^ is a very sensitive method for identifying genetic associations, it is essential that the groups being analysed are ethnically closely matched to avoid introducing bias due to non-disease-related patient stratification. To achieve this, the cases and controls were co-analysed with HapMap data using Taxonomy3^®^. Appropriately matched case and control populations for the trait being studied were selected using probabilistically defined regions using the Mahalanobis distance around centres determined by k-means clusters for each population. An appropriate α percentile was selected in order to retain the most subjects while minimising population heterogeneity. This approach is used to remove – if necessary – subjects causing biases in the analysis, and potentially confounding the case/control distinction.

#### Genetic ancestry

We carried out a co-analysis of the HapMap data with the RA (cases) and NBS (healthy controls) datasets, looking to define an ethnically homogenous sub-group of subjects. The objective of the analysis was to position RA and NBS subjects within a Caucasian/non-Caucasian ethnic contrast and to define an ethnically homogenous subgroup of Caucasians. The Taxonomy3^®^ analysis was conducted as follows:

– Reference population: HapMap Caucasians (CEU)
– Contrast populations: HapMap Chinese (CHB), Japanese (JPT), Tuscan (TSI) and Africans (YRI)
– Unknown subjects: RA and NBS subjects

### SNP-to-Gene Mapping

A custom SNP-to-gene mapping pipeline was used, which implemented both FUMA ^19^ and variant to gene (V2G) from Open Targets Genetics ^20^ to gather evidence. First, SNPs in high LD (r^2^ ≥ 0.6) with Taxonomy3^®^ SNPs were extracted from the EUR population of 1000 Genomes Data Phase 3 ^21^. These SNPs were then mapped to genes (“Tax3 genes”) by genomic distance (<40kb from gene boundary + 1kb promoter), predicted functional consequences (variant effect prediction – VEP ^22^), significant cis e/pQTLs (Open Targets Genetics collection) in relevant tissues, and chromatin mapping (Open Targets Genetics collection). These measures were then weighted and used to calculate a prioritisation score TOPSIS from the MCDA R package ^23^ based on a theoretical best and worst SNP-to-gene mapping. Standard evidence weights were based on Open Targets Genetics V2G scoring (functional prediction: 0.35, QTL evidence: 0.35, genomic distance: 0.2, chromatin proximity: 0.1). A TOPSIS score threshold of 0.4 was implemented to represent high quality mappings, whereby the SNP occurs within a gene or has a significant eQTL. This pipeline is available on GitHub (github.com/c4x-discovery/tax3-s2g).

### Assessment of Novelty

To assess the novelty of the findings at the SNP and gene level, data curated by the Open Targets ^24^ and Open Targets Genetics ^20^ platforms were used. All reported GWAS results related to EFO_0000685 (rheumatoid arthritis) in European populations were downloaded using the Open Targets Genetics API (November 2021) and lead SNPs were subjected to our SNP-to-gene mapping pipeline. Open Targets Indirect Disease Association Scores for Tax3 genes were investigated for novelty at the gene level (Open Targets download version 06.21).

### Bioinformatic Assessment of Drug Tractability

Target tractability details were downloaded from Open Targets ^24^ (Open Targets download version 11.21) to identify potential modalities to drug genetic targets directly or via relevant biological interactions. Scores were allocated to terms to aid visualisation of druggable targets. (Approved Drug: 1, Advanced Clinical: 0.9, Phase 1 Clinical: 0.8, Structure with Ligand: 0.5, UniProt loc high conf: 0.5, Literature: 0.5, GO CC high conf: 0.45, High-Quality Ligand: 0.4, UniProt loc med conf: 0.4, UniProt Ubiquitination: 0.4, High-Quality Pocket: 0.3, UniProt SigP or TMHMM: 0.3, Database Ubiquitination: 0.3, Med-Quality Pocket: 0.2, GO CC med conf: 0.2, Half-life Data: 0.2, Druggable Family: 0.1, Human Protein Atlas loc: 0.1, Small Molecule Binder: 0.1)

### Network and Pathway Enrichment

Network expansion was conducted on the protein-coding Tax3 genes from SNP-to-gene mapping, using experimental protein-protein interaction data from IntAct ^25^ (downloaded from Open Targets). A minimum IntAct score threshold of 0.5 was used to identify medium-high confidence known protein-protein interactions between Tax3 genes, and interactors were included that interacted with at least 2 Tax3 genes. This provided a network of potential interactors that may be occurring in relevant cells and tissues. Disease-relevant gene expression data was obtained from Genevestigator ^26^. 511 mRNA-seq samples were retrieved from studies relevant to RA, including only human samples labelled as RA or healthy control. This relevant set included blood and synovial tissues and cell types. Gene expression-based clustering was performed on Tax3 genes + interactors using WGCNA^27^ and clusters were analysed in the context of the protein-protein interaction network. Network and pathway enrichment was performed using the anRichment R package ^27^ using the built-in “GO” and “biosys” collections. Network visualisation and clustering was performed within R using igraph (Csardi and Nepusz, 2006) and <u>visNetwork</u>.

### Cell Assays

Preliminary validation of putative targets was performed using tool compounds in a cytokine release assay with peripheral blood mononuclear cells (PBMCs). Briefly, human PBMCs from healthy donors (n=3) were prepared from buffy coats and resuspended in RPMI-1640 containing 10% FBS, 1% penicillin/streptomycin, 2 mM L-glutamine and 50 μM 2-Mercaptoethanol. 1×10^5^ cells were added per well to a 96-well flat-bottomed plate, and for stimulated wells, anti-CD3 (final concentration 0.25 μg/mL) was added to cells immediately prior to seeding to the plate. Compounds (BML-210, CTLA-4 Fc Chimera, C4X_17358 and Merimepodib) were solubilised in DMSO with the final vehicle concentration in wells of 0.1%. Treatments were added in triplicate to wells in a final volume of 100 μL per well. Cells were cultured for 72 hours at 37°C, 5% CO_2_. At the end of the culture period, cells were stained for viability using eBioscience Fixable Viability Dye eFluor 780, and supernatants were collected for subsequent assessment of cytokine production by multiplex using ThermoFisher custom ProcartaPlex kits.

## Results

### Univariate Quality Control

Subjects and patients: Gender discrepancies were discovered in 19 subjects, 2 subjects had a missing value rate above 5%, and 5 subjects were found to be related: these subjects were removed from the analysis. A total of 26 subjects, predominantly RA patients, were removed due to univariate QC deviations.

Variables: The following steps were combined to establish the variables that were used in subsequent analyses (Table 1):

**Table 1.** Univariate QC procedures for SNPs (using the 90% genotype call probability threshold)

| Filter | Number of variables* | Outcome |
| --- | --- | --- |
| Monomorphic SNPs | 15,858 | Removed |
| > 5% missing value rate | 17,485 | Removed |
| SNPs departing from HWE in controls | 3,663 | Removed |

\* There are some overlaps between these filters (e.g. a SNP may be monomorphic and have >5% missing data)

We obtained the latest annotations file from Affymetrix pertaining to their discontinued 500K chip. Table 1 in supplementary material shows descriptive statistics for merged NBS & RA datasets and Table 2 in supplementary material shows the chromosomal location of available variables.

**Table 2.** Final Case/Control Cohorts

|  | Cases | Controls |
| --- | --- | --- |
| pre univariate QC | 1860* | 1480 |
| post univariate QC | 1834 | 1471 |
| post multivariate QC | 1563 | 1212 |
| post multivariate QC gender balanced | 793 | 1212 |

\*1860 subjects available from the initial 1999 after applying manufacturer's exclusion criteria.

From an initial list of 500,306 variables, the final QC’d data for multivariate QC analysis included 480,785 variables.

### Multivariate Quality Control

Chiamo generated genotype calls were used. A total of 480,785 SNPs shared by all datasets, non-monomorphic and having a total missing value rate lower than 5%, were analysed.

Fig. 1a shows the global Caucasian/non-Caucasian ethnic contrast. Subjects were somewhat dispersed, but the majority overlapped with the CEU cluster.

**Fig. 1.**
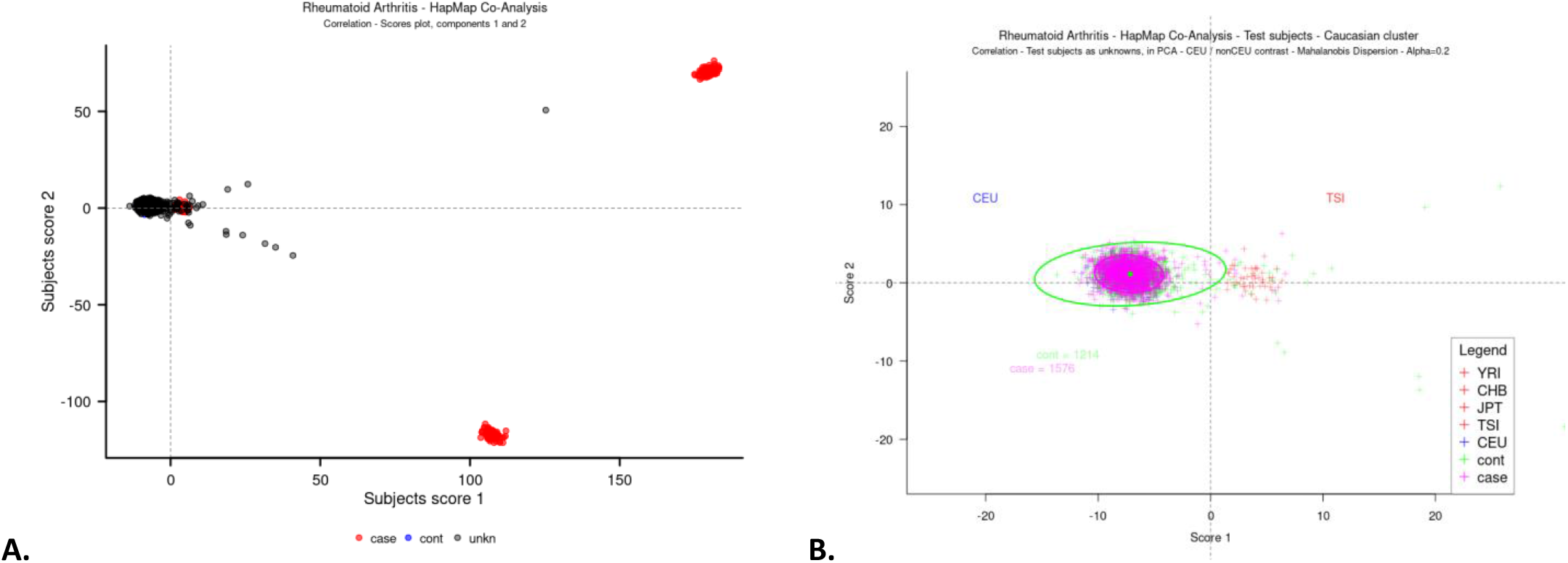
PCA score plot of Taxonomy3^®^ co-analysis of HapMap, RA (case) and NBS (control) subjects. **A** Caucasian/non-Caucasian ethnic contrast with RA (case), NBS (control) and other genetic ancestries as unknowns – in correlation PCA. **B** PCA score plot of Taxonomy3^®^ co-analysis of HapMap subjects. Patients within pink and green ellipses represent subjects associated with RA and NBS cohorts respectively, close to the CEU cohort. This is an expanded view of the region of interest.

Fig. 1b shows the ethnic boundaries for cases and controls defined using a Mahalanobis distance from the Caucasian cluster centre. A percentile value of α=0.2 was selected to define subjects in both cohorts using their respective ellipses.

The RA population in the WTCCC dataset was heavily skewed towards females. To reduce the chance for spurious associations the sample was gender balanced to achieve an odds ratio of 1.

The output from the multivariate analysis gives a final population of 2005 subjects (Table 2) Subsequent Taxonomy3^®^ analyses were restricted to these subjects.

### Taxonomy3^®^ Analysis Results

The primary output of Taxonomy3^®^ analysis is shown in the biplot in Fig. 2. The cases and controls are well separated by the 1^st^ principal component (PC), and the case/control dummy variable is closely aligned with the X-axis, showing that case/control separation is the biggest source of variation in the dataset (Fig. 2a). This is confirmed by the Scree plot (Fig. 2b) which shows that most variation is accounted for by the first principal component, with much smaller contributions from the other components. Variables projecting from the central origin along the case/control axis are relevant to discriminating cases and controls. 2^nd^ PC reveals a clustering pattern splitting both cases and controls into three distinct subgroups. Fig. 2c and d show the loci driving the separation in PC1 and PC2, respectively.

**Fig. 2.**
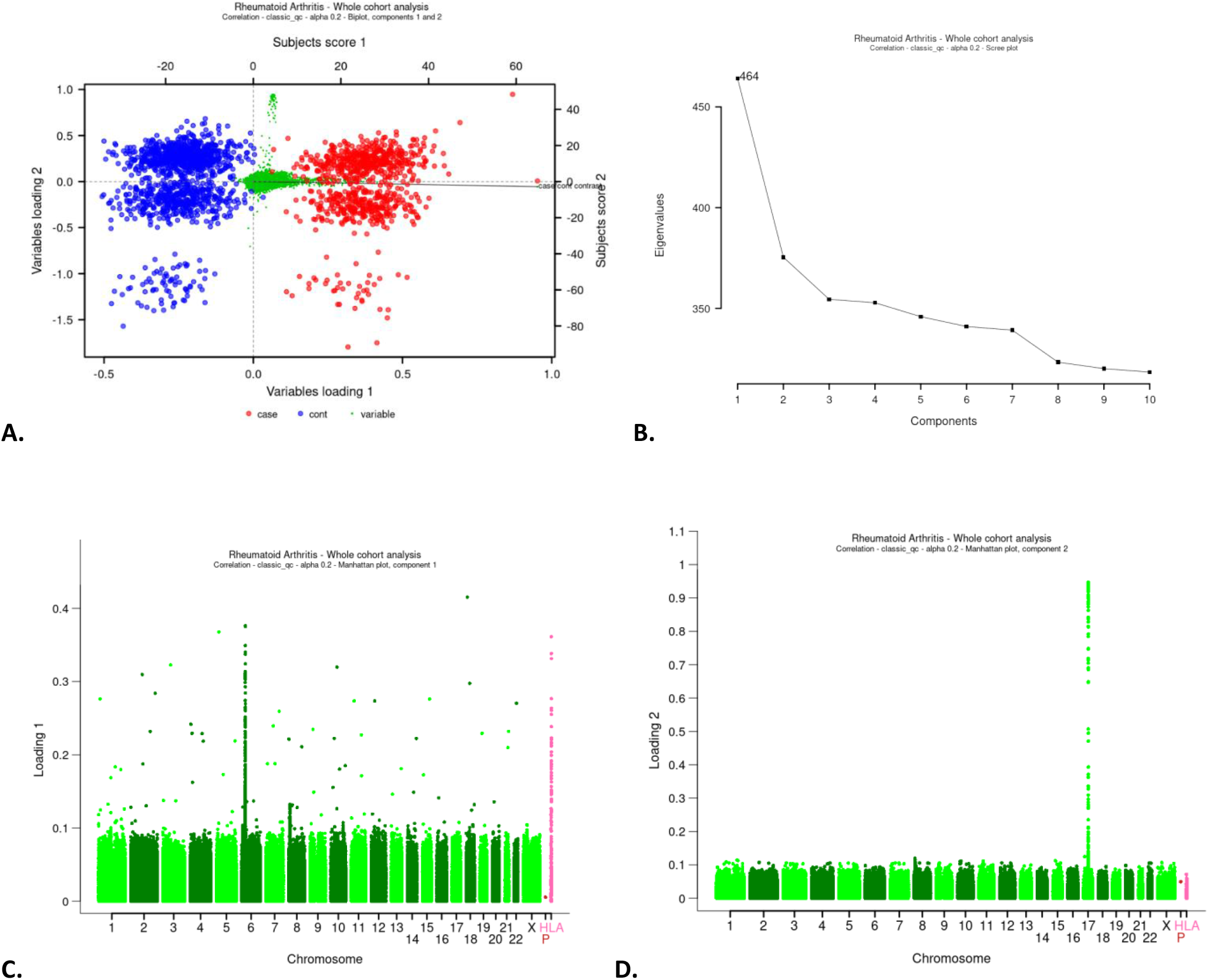
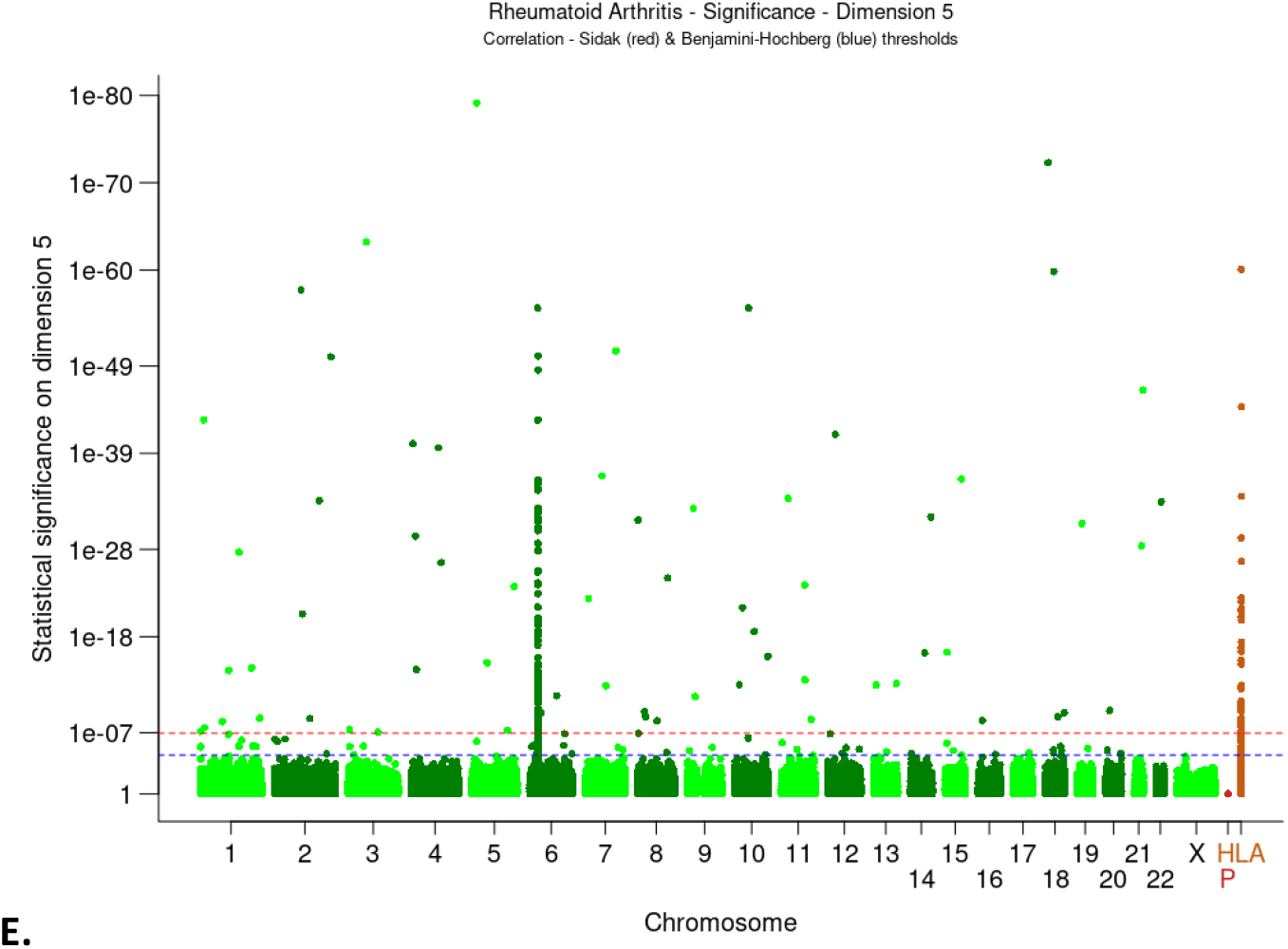
Taxonomy3^®^ analysis plots. **A** Biplot of the first two PCA components, showing NBS controls (blue), RA cases (red) and SNPs variables (green). **B** PCA Scree plot, showing that most of the signal was retained by the first few principal components. **C, D** Manhattan plots for loadings 1 and 2. **E** Manhattan plot based on 10, 000 permutations of the case/control status representing projected loadings as p-values. The red and blue dotted lines represent the Šidák and Benjamini-Hochberg whole genome statistical thresholds, respectively. Plotted as HLA are imputed HLA variables and as P -data collection centres

Permutation analysis was used to determine statistical significance of these findings. A total of 10,000 permutations of case/control labels and re-analysis were carried out. Based on the inspection of the Scree plot, 5 PCA components were retained. The Šidák genome wide significance threshold was used (alpha=5%, p-value = 1.13e-07). The resulting Manhattan plot is shown in Fig. 2e. Statistically significant loci were spread across the genome with large LD block located on chromosome 6. Many of the loci on chromosome 6 are located within the major histocompatibility complex (MHC) region as indicated by HLA imputed variables (on the Manhattan plot Fig. 3e plotted as chromosome HLA).

**Fig. 3.**
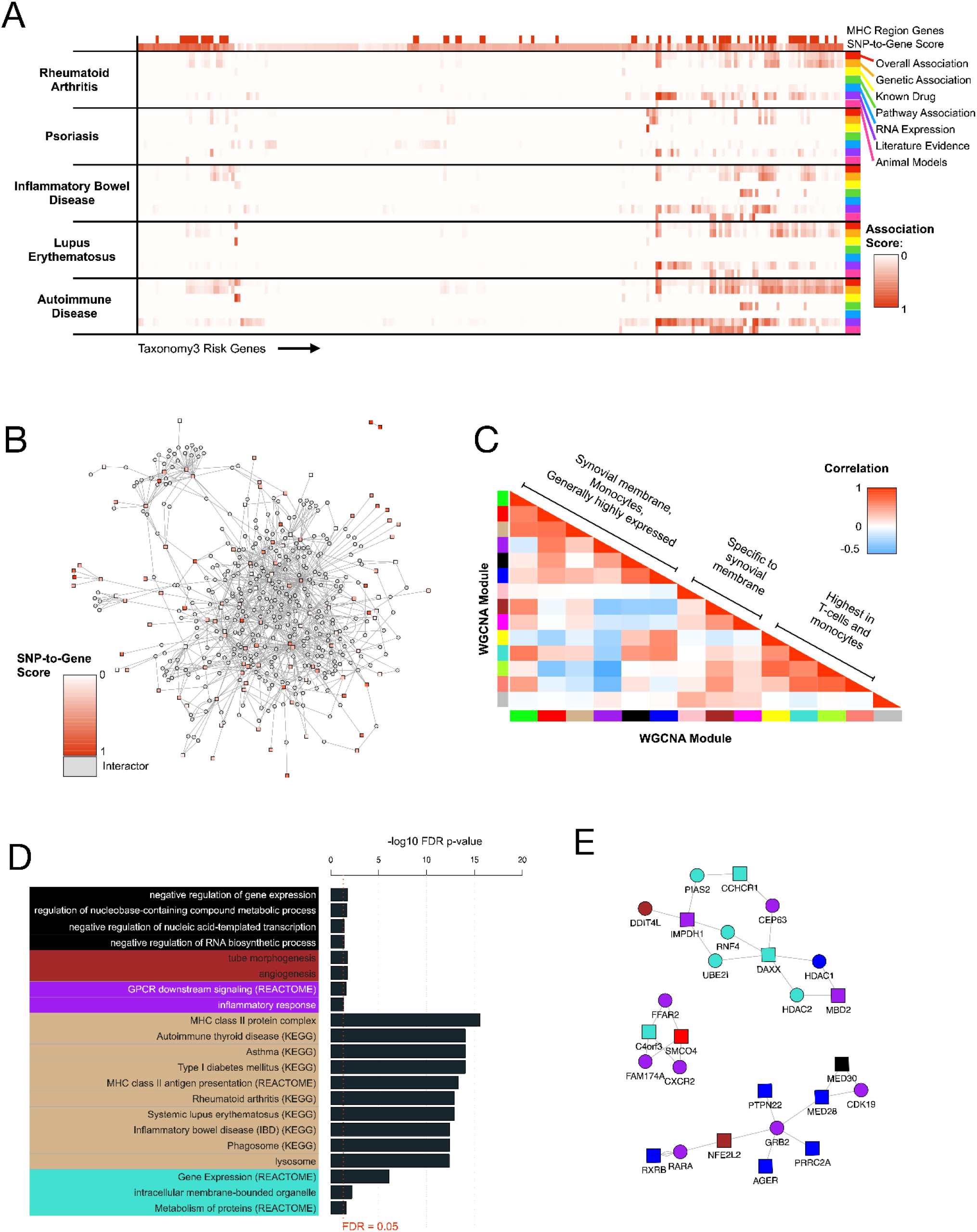
In-silico Exploration of Taxonomy3^®^ Findings. **A** Known disease associations with genes mapped from Tax3 SNPs, using data from the Open Targets platform. **B** Network of protein-protein interactions between Tax3 genes (red/pink) and known interactors, using data from the IntAct database. **C** Broad clustering of WGCNA co-expression modules, showing enrichment across various tissues relevant to RA. **D** Significantly enriched terms for WGCNA co-expression modules. **E** Network expansion of purple module genes. Square nodes represent Tax3 genes and circular nodes represent interactors

### Interpretation of Taxonomy3^®^ Findings (SNP-to-gene)

Taxonomy3^®^ genetic findings were mapped to genes using the parameters and pipeline described in Methods. The combined Taxonomy3^®^ analysis revealed 173 SNPs that exceeded the Šidák significance threshold. As expected, and consistent with variants detected by conventional analysis all of these SNPs except for one SNP (rs1800416) are located in non-coding regions of the genome. 109 significant SNPs mapped to the MHC region on chr6. The MHC region presents a challenge for SNP-to-gene mapping, as it is highly polymorphic, has a very high gene density and a low recombination rate resulting in a strong LD structure. Using the SNP-to-gene thresholds detailed in methods, a list of 233 genes was obtained (190 protein-coding genes).

### Novelty of Taxonomy3^®^ Findings

The novelty of Tax3 SNPs were assessed in two ways, by colocalising Taxonomy3^®^ genetic signals with published GWAS results, and using gene-level disease association scores from Open Targets ^24^ to explore known associations (genetic and others) between Tax3 genes and RA and related autoimmune disorders.

First, Tax3 SNPs were compared to significant GWAS results by matching exact rsIDs or matching rsIDs to SNPs in high LD (r^2^ ≥ 0.6 in 1KG Ph3 EUR). Using this method, 2 loci within the MHC region were matched (rs6457620/HLA-DQB1 and rs9268557/HLA-DRA) and 1 locus on chr1 (rs6679677/PTPN22). To expand this genetic mapping, significant GWAS SNPs were mapped to genes using the same pipeline as Tax3 SNPs. This identified 6 additional loci outside the MHC region mapping to the same top genes, RTN4IP1 and SUPT3H on chr6, ACOXL on chr2, TTC34 on chr1, LYZL1 on chr10 and AMOTL1 on chr11.

For gene-level disease association, 3 related autoimmune disorders were selected (psoriasis, inflammatory bowel disease (IBD) and systemic lupus erythematosus) based on their potential overlap of disease aetiology with RA and the understanding of heritable elements for these diseases. The scores are hierarchically cumulative, so the *autoimmune disease* scores represent an accumulated score for all autoimmune disorders according to Open Targets ^24^. Fig. 3a displays a heatmap of these scores, excluding 86 Tax3 genes that have a value of 0 for all selected scores. SNP-to-gene evidence (TOPSIS) is also displayed, and MHC region genes are labelled. Rows and columns are hierarchically clustered using hclust from the fastcluster ^29^ R package. This analysis revealed 3 additional weak known genetic associations with RA (chr2 rs4338920 LRP1B, chr13 rs17086772 SLC46A3 and chr4 rs17669915 LEF1). These weak associations are from GWAS for non-European populations, case-case comparative GWAS and similar disease GWAS. Other interesting genes without known genetic association with RA include 2 genes targeted by drugs in RA clinical trials (PDE5A: Dipyridamole, IMPDH1: Mizoribine), 1 additional gene targeted by drugs in clinical trials for other autoimmune diseases (ADRA1: Isoxsuprine for multiple sclerosis), 6 genes with known genetic association to autoimmune diseases but not RA (C1QTNF6, BCL2L11, KAZN, ETV3, MACROD2, PROK2, KCNE4), and 3 genes from pathways linked to IBD (PROK2, CYTH4, RAC2).

Taxonomy3^®^ findings have been shown to support many existing genetic associations with RA, reveal unknown genetic associations with genes that are known to be associated with RA or related autoimmune disorders, and provide 150 novel, genetically associated RA targets for further validation and exploration.

### *In-silico* exploration of Taxonomy3^®^ Findings

Bioinformatics analysis was performed to interpret and prioritise the Tax3 genes. The 190 protein-coding genes underwent enrichment analysis as described in Methods. The top significantly enriched terms included Antigen processing and presentation (KEGG) (FDR p=3.96e-03), MHC class II antigen presentation (REACTOME) (FDR p=8.27e-03), and many KEGG terms for autoimmune and infectious diseases, including Staphylococcus aureus infection (FDR p=3.96e-03), Autoimmune thyroid disease (FDR p=8.27e-03), Systemic lupus erythematosus (FDR p=1.69e-02), Asthma (FDR p=8.27e-03), and Rheumatoid arthritis (p=1.69e-02). These results were highly dominated with HLA genes, so a separate enrichment was performed excluding genes from the MHC region. The top terms for this analysis were transferase activity (p=1.53e-03) and bone morphogenic protein (BMP) signalling pathway (p=1.53e-03), the latter of which has been linked to autoimmune disease ^30^.

Genetic association can only elucidate part of the mechanistic network for complex diseases, and our inclusive SNP-to-gene mapping pipeline will include false positives, so in order to contextualise our gene list and encourage dropout of false positives, a network expansion and clustering approach was undertaken. Experimentally validated protein-protein interactions (PPI) from IntAct ^25^ were obtained between genes within the 190 protein-coding Tax3 genes, and also between interactor proteins and at least 2 Tax3 genes. The resulting PPI network included 114 Tax3 genes and 359 interactors. Network visualisation and clustering revealed some peripheral clusters, some clusters formed of pairs of Tax3 nodes with many common interactors, and a large central subnetwork of interconnected nodes (Fig. 3b). These interactions are mainly from *in-vitro* studies, so represent potential protein-protein interactions that could occur in a physiological context. To interpret this network and identify relevant clusters and subnetworks, 2 complementary approaches were undertaken, PPI network community identification using network clustering methods within igraph ^28^, and network coexpression clustering using WGCNA ^27^.

PPI subnetworks were identified using the cluster walktrap method from the igraph R package ^28^. This identified 20 clusters with at least 5 members. For coexpression clustering, relevant gene expression data was downloaded from Genevestigator ^26^. 511 patient samples from RA studies were selected, representing multiple disease-relevant tissue types, and 233 Tax3 genes + 349 interactors underwent network coexpression clustering using WGCNA ^27^. This identified 14 clusters, 13 of which had distinct eigengene expression profiles in RA-relevant tissues. Correlation analysis of cluster eigengenes revealed 3 overall cluster groups (Fig. 3c) representing targets that were 1: widely expressed but highest in synovial membrane and monocytes, and low in fibroblast synoviocytes, 2: synovial membrane-specific, 3: highest expression in T-cells and monocytes, showing that Tax3 genes are enriched for genes expressed in highly disease-relevant cell types. 5 of these coexpression clusters had significantly enriched pathway terms, using a combined collection of GO, KEGG and REACTOME terms (Fig. 3d). The tan cluster was highly enriched for MHC class II genes, and many diseases for which MHC-II factors are associated. The purple cluster was enriched for inflammatory response genes (FDR p=4.53e-02). This cluster did not feature any direct protein-protein interactions, but expanding this cluster for direct interactors displayed multiple purple interactor genes that may provide additional druggable targets to perturb the RA-specific inflammatory network identified through Tax3 genetic associations (Fig. 3e). This analysis provides potential drug targets that could be taken forward into a variety of assays to explore and validate their role in disease aetiology.

Small-molecule druggability (tractability) assessment was also conducted on network nodes using data from the Open Targets platform ^24^ as described in methods. 3 additional interactor genes were identified that are targeted by drugs in clinical trials for autoimmune diseases (FGFR3: Masitinib for RA, KCNA3: Dalfampridine for multiple sclerosis, PSMB5: Bortezomib and Ixazomib for autoimmune thrombocytopenic purpura) In order to progress the analysis and validate potential therapeutic targets, genes with available tool compounds were selected that may impact and disrupt the disease network.

### In vitro validation of Taxonomy3^®^ findings

In an attempt to validate the novel genetic associations from Taxonomy3^®^ analysis in a preliminary cellular context, we identified those putative RA targets which were entirely novel, and those established RA targets without a genetic link that have been previously found to be expressed in leukocytes. Publicly available tool compounds were available for MEF2B and IMPDH1, and an internal compound (C4X_17358) was synthesized as an inhibitor of the potassium channel Kv1.3 (IC_50_ 2nM determined via SyncroPatch, data not shown), of which the Tax3 gene KCNE4 is a functional co-factor (Table 3). These compounds were tested in the PBMC validation assay as described in the Methods. CTLA-4 Fc Chimera was included in the study as a positive control ^31,32^. Whilst it not certain that the putative genetic variants modulate the mapped genes in PBMCs specifically, using tool compounds to investigate potential modulation of inflammatory pathways in these cells would provide justification to investigate this further.

**Table 3.**
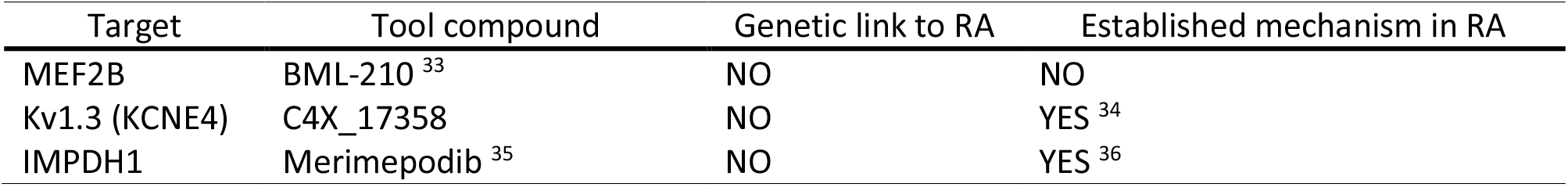
Taxonomy3^®^ targets to be tested in PBMC validation assay

| Target | Tool compound | Genetic link to RA | Established mechanism in RA |
| --- | --- | --- | --- |
| MEF2B | BML-210 <sup>33</sup> | NO | NO |
| Kv1.3 (KCNE4) | C4X_17358 | NO | YES <sup>34</sup> |
| IMPDH1 | Merimepodib <sup>35</sup> | NO | YES <sup>36</sup> |

Following 72 hours of treatment, no compound was found to reduce cell viability below 75% (Fig. 4). Pro-inflammatory cytokine measurements were carried out for (interleukins) IL-17a, IL-6, interferon gamma (IFNγ) and tumour necrosis factor alpha (TNFα) (Fig. 4a-d) in cells stimulated with anti-CD3. BML-210 treatment resulted in a trend to further increase inflammatory cytokine release with increasing compound concentration (Fig. 4a). Comparatively, treatment with C4X_17358, merimepodib or CTLA-4 Fc Chimera resulted in a trend toward a decrease in pro-inflammatory cytokines with increasing compound concentration (Fig. 4b-d).

**Fig. 4.**
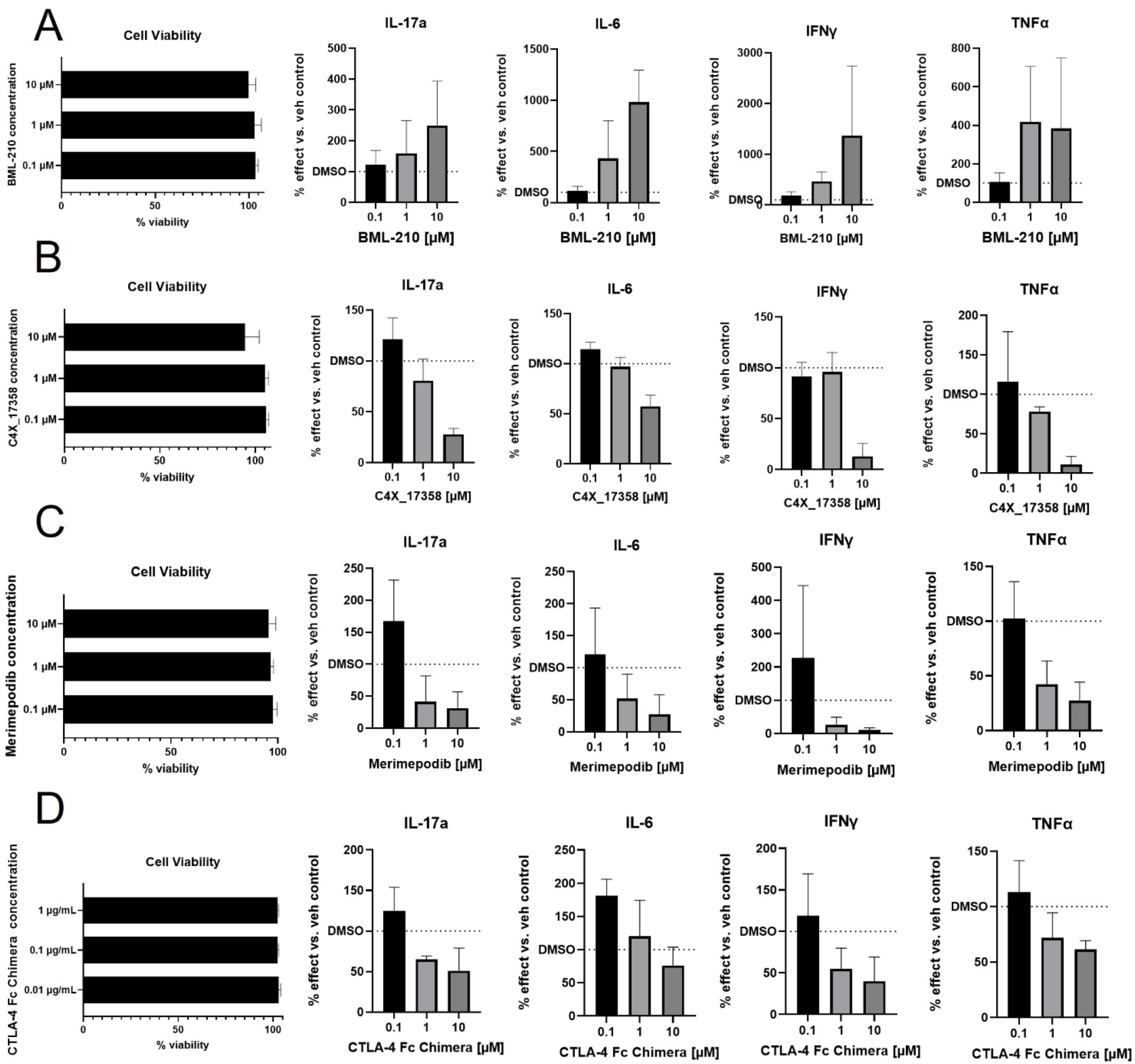
PBMC assay validation of Taxonomy3^®^ targets. PBMCs were cultured, stimulated with anti-CD3 and treated with tool compounds as listed for 72 hours. **A** BML-210 treatment. **B** C4X_17358 treatment. **C** Merimepodib treatment. **D** CTLA-4 Fc Chimera treatment. Viability of PBMCs was measured via eBioscience Fixable Viability Dye eFluor 780. Viability is represented as a percent of the viability of vehicle control cells. Levels of secreted IL-17a, IL-6, IFNγ and TNFα were determined via multiplex. Levels of cytokines are represented as a percent of cytokine secreted compared to vehicle control cells. PBMC assays were performed in technical triplicates, and an average of the three values taken to represent one biological replicate; three biological replicates were performed (three different donors)

## Discussion

To date, divergences – which were originally conceived in vector calculus to explore fluid mechanics -have not been extensively used in biology. LBFs have been used in assessing diagnostic kit performance ^37^, and more recently they have been used in Bayesian Analysis of Gene Essentiality (BAGEL) methodology to analyse pooled CRISPR studies ^38^. Taxonomy3^®^ is the first application of which we are aware that applies individualised LBFs to human genetic data linked to a binary outcome. By replacing individual genotypes with the corresponding LBF, a wide range of linear algebra methods can be brought to bear on the genetic data. Analysing the LBF matrix using PCA generates a number of outputs:

- Variables of interest relevant for case/control discrimination;
- Heterogeneity in the case/control populations, allowing for sub-groups within the populations to be identified;
- Variables of interest relevant for sub-group separation.

As this is a different approach to exploring genetic data, it is not surprising that the outputs are not identical to the output of the conventional GWAS analysis. In the original analysis of this RA case/control dataset ^11^, two peaks were seen, one on chromosome 6 in the MHC region, and one on chromosome 1. The Manhattan plot from our Taxonomy3^®^ analysis (Fig. 2 a, e) shows that we have replicated these two findings and also greatly increased the number of significant associations. Comparison of our findings with the data currently available on Open Targets shows that while some of our additional findings have been observed in other, subsequent larger studies or meta-analyses (which provides additional validation for our methodology), there are other findings that provide truly novel insights.

Bioinformatic analysis of the Tax3 genes showed significant clustering in immune-related pathways (Fig. 3 b), providing good validation for the newly-identified genes. Likewise, co-expression clustering analysis showed that changes of expression of the genes were enriched in T-cells, synovial tissues and monocytes – highly disease-relevant cell types (Fig. 3 c).

Suitable tool compounds were only available for a few of the Tax3 genes – Kv1.3 (KCNE4), IMPDH1 and MEF2B: two of these are of particular note:

### KCNE4

Association with this gene was identified through Taxonomy3^®^ analysis. This gene has not previously been associated with RA, and codes for an accessory protein modulating the activity of Kv1.3 ^39,40^. This K^+^ channel is important for the functioning of TEM cells, which have a key role in maintaining the autoimmune drive in RA ^41–43^. For this reason, inhibiting this channel has been proposed as a possible target for various inflammatory diseases ^44–46^ including RA. We consider that the addition of genetic support for the pathway from our Taxonomy3^®^ analysis significantly increases the likelihood of successful clinical development for inhibitors of Kv1.3.

### MEF2B

This gene mapped from a SNP identified in Taxonomy3^®^ analysis. The gene has not been associated with RA in published GWAS, and there is limited data to implicate the gene in RA. Examination of the tool compound BML-210, that blocks the interaction of MEF2 with histone deacetylase (HDAC), in *in vitro* models showed consistent effects of inhibition of MEF2B on immune cell function. This result underlines the ability of Taxonomy3^®^ analysis to generate novel genetic insights, adding significantly to our knowledge of disease aetiology, and flagging novel drug targets with genetic support.

Association with NR4A3 was also identified in Taxonomy3^®^ analysis passing the Benjamini-Hochberg genome wide significance threshold (false discovery rate correction). This nuclear receptor is known to impact expression of FoxP3, the key gene required for regulatory T-cell (Treg) formation ^47–49^. A number of publications have demonstrated a dysfunction of Treg cells in RA patients ^50–52^, and therapeutic modulation of Tregs for RA has been proposed ^53^. Unfortunately, no suitable tool compounds were available to probe the functioning of this gene in the PBMC model.

There are a number of limitations to this study. Firstly, we have not been able to analyse other datasets (potentially from other genetic ancestries) to examine the translation of our findings to other populations. Secondly, as with all genetic studies, the inherent uncertainties of SNP-to-gene mapping means that the genes described here may not be the true genes involved in the genetic susceptibility detected by Taxonomy3^®^. However, we have maximised the chance of including the causal gene by using a combination of positional and functional mapping in our SNP-to-gene process and downstream triaging to identify the genes with a high probability of impacting disease aetiology. Furthermore, as non-coding variants are thought to regulate genes in a cell type specific manner, it is possible that some or all of the subset of Tax3 genes we examined in the PMBC assay (MEF2B, Kv1.3 (KCNE4), IMPDH1) are in fact regulated by the variants detected, in a different cell type (e.g. fibroblast synoviocytes). Once confidence in a SNP-gene mapping is achieved, along with demonstrating clear impact of the target in a disease relevant context (conduct of additional studies extending beyond the remit of this publication) validation of the genetics will need to be confirmed e.g. show differential regulation of the gene in question depending on which allele is present. As the genotypes for the PBMC donors were not available this could not be examined alongside the tool compound examination.

In this paper, we describe a unique method of analysing genetic datasets, which extracts additional genetic insights in a hypothesis-free way to complement those discovered through conventional analysis of GWAS. The results of our Taxonomy3^®^ analyses have significant value for drug discovery programmes for treating RA, the identified novel targets (e.g. MEF2B) benefitting from the increased chances of success due to their genetic association with the disease subject to further experimental validation. The method can be applied to all datasets with well-defined dichotomous cohorts e.g. case/control, mild/severe, responders/non-responders, and has the added benefit that other data types (e.g. mRNA expression, clinical data, longitudinal phenotypes) can all be converted into LBFs and co-analysed with genetic data ^15^, although this application of the method is currently limited by the availability of suitable datasets. The method can identify novel genes or pathways of interest for a disease (or other phenotype of interest) leading to innovative drug discovery programmes with the added confidence of genetic support.

#### Data Availability

This study makes use of data generated by the Wellcome Trust Case Control Consortium. A full list of the investigators who contributed to the generation of the data is available from www.wtccc.org.uk. Funding for the project was provided by the Wellcome Trust under award 076113. A Accession numbers for data used in this study: EGAD00000000007 -WTCCC1 project Rheumatoid arthritis (RA) samples, EGAD00010000250 -NBS control samples. Access to summary data and individual-level genotype data is available by application to the Wellcome Trust Case Control Consortium Data Access Committee.

## Data Availability

This study makes use of data generated by the Wellcome Trust Case Control Consortium. A full list of the investigators who contributed to the generation of the data is available from www.wtccc.org.uk. Funding for the project was provided by the Wellcome Trust under award 076113. A Accession numbers for data used in this study: EGAD00000000007 - WTCCC1 project Rheumatoid arthritis (RA) samples, EGAD00010000250 - NBS control samples. Access to summary data and individual-level genotype data is available by application to the Wellcome Trust Case Control Consortium Data Access Committee

## Acknowledgements

We thank Olivier Delrieu for designing the concept of the analysis method and creating the Taxonomy3^®^ analysis pipeline.

## Author Contributions

Design or the analysis, J.K.; interpretation of the results, J.K. AM, M.C., N.HK.; writing of the manuscript, J.K., A.M., N.HK., M.C. figure and table preparation, J.K, N.HK., M.C.; bioinformatics analysis, N.HK.; data analysis, A.P., N.HK., M.C.; collection and/or assembly of data, J.K.; Biology project management, A.S.M. ; final approval of the manuscript, C.M; all authors revised the manuscript.

## Statements and Declarations

### Conflict of interest

The authors declare no competing interests.

## Notes

### Competing Interest Statement

The authors have declared no competing interest.

### Funding Statement

This study did not receive any funding.

### Author Declarations

Access to summary data and individual-level genotype data is available by application to the Wellcome Trust Case Control Consortium Data Access Committee. No ethical approval/IRB review was required for this work.

