## Supplemental Material for "New Genetic Insights in Rheumatoid Arthritis using Taxonomy3^®^, a Novel method for Analysing Human Genetic Data"

**Supplementary Materials:**

**Table 1	Descriptive stats for merged NBS & RA datasets**

| number of genotypes | 1,671,022,040 |
| --- | --- |
| number of variables | 500,306 |
| number of subjects | 3,340 |
| % Genotypes with call probability < 95% | 1.92 % |
| % Genotypes with call probability < 100% | 5.03 % |

**Table 2	Chromosomal location of available variables**

| **Location** | **Number of variables** |
| --- | --- |
| Autosomes | 490,032 |
| X | 10,274 |
| 1 | 40220 |
| 2 | 41400 |
| 3 | 33801 |
| 4 | 32334 |
| 5 | 32056 |
| 6 | 31470 |
| 7 | 25835 |
| 8 | 27457 |
| 9 | 22864 |
| 10 | 28501 |
| 11 | 26273 |
| 12 | 24954 |
| 13 | 19188 |
| 14 | 15721 |
| 15 | 14356 |
| 16 | 15309 |
| 17 | 11281 |
| 18 | 14881 |
| 19 | 6399 |
| 20 | 12400 |
| 21 | 7125 |
| 22 | 6207 |

**Table 3 Subjects part of the RA/HapMap co-analysis**

| **Collection** | **Cohort** | **Number of Subjects** |
| --- | --- | --- |
| HapMap | CEU (Caucasians) | 172 |
| HapMap | CHB (Chinese) | 44 |
| HapMap | JPT (Japanese) | 40 |
| HapMap | TSI (Tuscan) | 48 |
| HapMap | YRI (Africans) | 85 |
| WTCCC1 | NBS | 1473 |
| WTCCC1 | RA | 1860 |

**Table 4 Taxonomy3® Significant SNPs**

| variable | chromosome | p-value | gene |
| --- | --- | --- | --- |
| p80_G_HLA_DQA1_01 | 6 | 2.76547E-11 | HLA-DQA1 |
| p80_G_HLA_DQA1_01.02 | 6 | 4.85555E-13 | HLA-DQA1 |
| p80_G_HLA_DQA1_02 | 6 | 5.55972E-09 | HLA-DQA1 |
| p80_G_HLA_DQA1_03 | 6 | 8.39E-61 | HLA-DQA1 |
| p80_G_HLA_DQA1_03.01 | 6 | 4.85E-21 | HLA-DQA1 |
| p80_G_HLA_DQA1_03.03 | 6 | 4.93E-30 | HLA-DQA1 |
| p80_G_HLA_DQA1_05 | 6 | 7.07644E-11 | HLA-DQA1 |
| p80_G_HLA_DQB1_02 | 6 | 3.64752E-13 | HLA-DQB1 |
| p80_G_HLA_DQB1_03 | 6 | 4.57E-45 | HLA-DQB1 |
| p80_G_HLA_DQB1_03.01 | 6 | 5.20E-17 | HLA-DQB1 |
| p80_G_HLA_DQB1_03.02 | 6 | 2.37E-27 | HLA-DQB1 |
| p80_G_HLA_DQB1_06 | 6 | 6.43E-22 | HLA-DQB1 |
| p80_G_HLA_DQB1_06.02 | 6 | 1.37382E-08 | HLA-DQB1 |
| p80_G_HLA_DQB1_06.03 | 6 | 2.12079E-08 | HLA-DQB1 |
| p80_G_HLA_DRB1_04 | 6 | 3.80E-23 | HLA-DRB1 |
| p80_G_HLA_DRB1_04.01 | 6 | 1.29E-20 | HLA-DRB1 |
| p80_G_HLA_DRB1_13 | 6 | 2.95439E-11 | HLA-DRB1 |
| p80_G_HLA_DRB1_15 | 6 | 2.16839E-09 | HLA-DRB1 |
| p80_G_HLA_DRB1_15.01 | 6 | 1.7265E-10 | HLA-DRB1 |
| rs1018433 | 6 | 2.8164E-08 | C6orf10 |
| rs10212068 | 22 | 3.42E-34 | C1QTNF6 |
| rs10262109 | 7 | 1.89E-51 | PTPRZ1 |
| rs10499044 | 6 | 5.8631E-12 | QRSL1 |
| rs10501805 | 11 | 1.28E-24 | TAF1D |
| rs1051336 | 6 | 1.04199E-08 | HLA-DRA |
| rs10843660 | 12 | 6.84E-42 | IPO8 |
| rs10910099 | 1 | 7.23883E-08 | MMEL1 |
| rs10947378 | 6 | 1.01886E-09 | HLA-DPA1 |
| rs11671119 | 19 | 1.10E-31 | MEF2BNB |
| rs11998409 | 8 | 4.12917E-10 | TRIM35 |
| rs12201454 | 6 | 5.31E-32 | BTNL2 |
| rs1265777 | 6 | 1.92E-33 | C6orf10 |
| rs12670243 | 7 | 4.008E-13 | SEMA3E |
| rs13425033 | 2 | 1.88E-58 | FHL2 |
| rs1367731 | 6 | 1.10422E-07 | HLA-DOA |
| rs1369036 | 1 | 5.02947E-09 | LMO4 |
| rs1410707 | 9 | 6.90416E-12 | HMGB3P23 |
| rs1440065 | 2 | 8.93E-51 | ACSL3 |
| rs1553460 | 4 | 3.02E-30 | QDPR |
| rs1559873 | 6 | 1.00277E-08 | C6orf10 |
| rs1559874 | 6 | 1.34296E-09 | C6orf10 |
| rs160619 | 5 | 5.23919E-08 | No gene |
| rs16838195 | 1 | 2.07E-28 | ETV3L |
| rs16898046 | 5 | 1.02208E-15 | ZSWIM6 |
| rs16908561 | 9 | 1.94E-33 | IZUMO3 |
| rs16916476 | 11 | 1.45E-34 | BBOX1 |
| rs16942813 | 15 | 8.79E-37 | AKAP13 |
| rs16957658 | 18 | 1.47248E-09 | C18orf54 |
| rs16968559 | 16 | 3.94909E-09 | XYLT1 |
| rs17012953 | 1 | 3.87546E-15 | RP11-565N2.1 |
| rs17050351 | 4 | 3.28E-27 | PDE5A |
| rs17086772 | 13 | 3.54214E-13 | POMP |
| rs17100135 | 14 | 1.96E-32 | RTL1 |
| rs17104722 | 14 | 7.17E-17 | VASH1 |
| rs17166781 | 7 | 4.30E-23 | ARL4A |
| rs17421624 | 6 | 1.14E-24 | TNXB |
| rs17665418 | 3 | 6.06E-64 | PROK2 |
| rs17669915 | 4 | 2.27E-40 | LEF1 |
| rs1785863 | 11 | 8.48149E-14 | AMOTL1 |
| rs1903736 | 3 | 8.61794E-08 | LSAMP |
| rs1948674 | 8 | 1.83E-25 | MED30 |
| rs2001097 | 6 | 5.37363E-08 | HLA-DRA |
| rs2001099 | 6 | 8.43439E-08 | HLA-DRA |
| rs206015 | 6 | 4.34453E-11 | NOTCH4 |
| rs2072633 | 6 | 2.49E-16 | CFB |
| rs2075800 | 6 | 1.15E-23 | HSPA1L |
| rs2076530 | 6 | 2.14E-33 | BTNL2 |
| rs2076533 | 6 | 2.72E-36 | BTNL2 |
| rs2121526 | 10 | 2.23E-56 | PCDH15 |
| rs2213580 | 6 | 7.92066E-08 | HLA-DRA |
| rs2227127 | 6 | 6.12132E-08 | HLA-DQA2 |
| rs2395161 | 6 | 8.65086E-08 | HLA-DRA |
| rs2395164 | 6 | 3.38036E-08 | HLA-DRA |
| rs2395167 | 6 | 3.1464E-08 | HLA-DRA |
| rs241427 | 6 | 2.36239E-09 | TAP2 |
| rs2497828 | 10 | 3.14604E-13 | SLC39A12 |
| rs2647046 | 6 | 2.99E-31 | HLA-DRB1 |
| rs2736172 | 6 | 5.06769E-13 | PRRC2A |
| rs2789336 | 1 | 2.57962E-08 | PAX7 |
| rs2838846 | 21 | 5.73E-47 | POFUT2 |
| rs2856816 | 6 | 2.83123E-09 | HLA-DPA1 |
| rs2857697 | 6 | 1.8619E-12 | PRRC2A |
| rs2859090 | 6 | 4.96036E-08 | HLA-DQA2 |
| rs2894249 | 6 | 8.12372E-13 | C6orf10 |
| rs2941794 | 18 | 5.00711E-10 | SALL3 |
| rs2943570 | 8 | 4.37731E-09 | HNF4G |
| rs3115560 | 6 | 1.08191E-09 | C6orf10 |
| rs3115576 | 6 | 2.10E-19 | C6orf10 |
| rs3122348 | 10 | 4.63E-22 | LYZL2 |
| rs3128947 | 6 | 1.78525E-15 | HLA-DPA1 |
| rs3129768 | 6 | 2.83762E-12 | HLA-DRA |
| rs3129900 | 6 | 4.22133E-08 | HLA-DRA |
| rs3129932 | 6 | 2.3712E-13 | C6orf10 |
| rs3129934 | 6 | 2.86516E-08 | HLA-DRA |
| rs3130311 | 6 | 8.60E-19 | C6orf10 |
| rs3132928 | 6 | 9.16791E-10 | C6orf10 |
| rs3132959 | 6 | 8.44698E-15 | C6orf10 |
| rs3134926 | 6 | 1.84606E-10 | NOTCH4 |
| rs3135363 | 6 | 5.40782E-12 | HLA-DOB |
| rs3135366 | 6 | 5.57487E-08 | HLA-DRA |
| rs3135376 | 6 | 7.05777E-08 | HLA-DRA |
| rs3135377 | 6 | 4.86653E-14 | C6orf10 |
| rs3135378 | 6 | 4.29518E-08 | HLA-DRA |
| rs3135393 | 6 | 8.82174E-09 | HLA-DRA |
| rs370233 | 18 | 5.06E-73 | PIEZO2 |
| rs3763307 | 6 | 3.37E-32 | BTNL2 |
| rs3806156 | 6 | 7.04E-25 | BTNL2 |
| rs3821009 | 2 | 2.65E-34 | PDE11A |
| rs3916765 | 6 | 1.51937E-12 | HLA-DQA2 |
| rs3957146 | 6 | 2.52E-26 | HLA-DQB1 |
| rs424232 | 6 | 4.44867E-08 | HLA-DQA2 |
| rs4338920 | 2 | 2.36816E-09 | LRP1B |
| rs4428528 | 6 | 2.62778E-11 | HLA-DRA |
| rs4697298 | 4 | 5.7803E-15 | GPR125 |
| rs4718582 | 7 | 3.73E-37 | TYW1 |
| rs4799934 | 18 | 1.46E-60 | CELF4 |
| rs486416 | 6 | 5.14735E-08 | EHMT2 |
| rs4959053 | 6 | 2.30E-56 | PSORS1C1 |
| rs4959093 | 6 | 7.06E-32 | C6orf10 |
| rs539002 | 6 | 5.02679E-10 | SUPT3H |
| rs539703 | 6 | 5.17E-33 | C6orf10 |
| rs574710 | 6 | 3.63E-33 | C6orf10 |
| rs6131746 | 20 | 2.9169E-10 | MACROD2 |
| rs615672 | 6 | 2.67712E-15 | HLA-DRB1 |
| rs644045 | 6 | 1.89839E-12 | CFB |
| rs6451916 | 5 | 7.24E-80 | RP11-774D14.1 |
| rs6457617 | 6 | 2.78E-49 | HLA-DQB1 |
| rs6457620 | 6 | 6.98E-51 | HLA-DQB1 |
| rs6679677 | 1 | 7.33665E-15 | PTPN22 |
| rs6701541 | 1 | 2.1161E-09 | FMN2 |
| rs692775 | 11 | 2.91387E-09 | NECTIN1 |
| rs6930933 | 6 | 3.33431E-08 | HLA-DRA |
| rs6936204 | 6 | 2.80E-18 | NOTCH4 |
| rs6992071 | 8 | 1.52352E-09 | NRG1 |
| rs707939 | 6 | 4.25E-26 | HSPA1L |
| rs707974 | 6 | 5.71E-19 | TNXB |
| rs7080464 | 10 | 1.84E-16 | PPP2R2D |
| rs7194 | 6 | 2.12E-29 | HLA-DRA |
| rs733208 | 6 | 3.3162E-09 | HLA-DPB1 |
| rs7539166 | 1 | 1.45E-43 | TMEM51 |
| rs7632506 | 3 | 4.69581E-08 | EDEM1 |
| rs7694697 | 4 | 7.50E-41 | SORCS2 |
| rs7746922 | 6 | 1.51865E-13 | HLA-DRA |
| rs7747521 | 6 | 2.77318E-14 | HLA-DRA |
| rs7749092 | 6 | 1.23649E-15 | HLA-DQA2 |
| rs7756262 | 6 | 7.64E-31 | HLA-DRA |
| rs7766843 | 6 | 1.22849E-13 | HLA-DRA |
| rs7767325 | 6 | 2.05983E-09 | C6orf10 |
| rs7775228 | 6 | 4.34972E-11 | HLA-DQB1 |
| rs7818865 | 8 | 4.35E-32 | ERICH1 |
| rs7996435 | 13 | 2.45557E-13 | IRS2 |
| rs805297 | 6 | 1.03E-17 | PRRC2A |
| rs8129909 | 21 | 3.97E-29 | IGSF5 |
| rs899848 | 15 | 5.98E-17 | TJP1 |
| rs910049 | 6 | 6.49282E-14 | C6orf10 |
| rs910050 | 6 | 7.15855E-08 | C6orf10 |
| rs926591 | 6 | 6.30E-33 | C6orf10 |
| rs9267954 | 6 | 1.03E-35 | NOTCH4 |
| rs9267971 | 6 | 3.37E-19 | C6orf10 |
| rs9268207 | 6 | 6.73266E-09 | C6orf10 |
| rs9268230 | 6 | 8.13366E-10 | HLA-DQA2 |
| rs9268402 | 6 | 2.21E-32 | BTNL2 |
| rs9268403 | 6 | 3.91E-31 | BTNL2 |
| rs9268429 | 6 | 1.93E-29 | BTNL2 |
| rs9268480 | 6 | 5.01E-31 | BTNL2 |
| rs9268557 | 6 | 2.18E-19 | HLA-DRA |
| rs9268560 | 6 | 1.57E-43 | HLA-DRA |
| rs9268645 | 6 | 1.05E-36 | HLA-DRA |
| rs9268831 | 6 | 1.46599E-11 | HLA-DRA |
| rs9268853 | 6 | 2.02675E-15 | HLA-DQA1 |
| rs9268856 | 6 | 1.11758E-13 | HLA-DRA |
| rs9268858 | 6 | 1.51E-28 | HLA-DQA1 |
| rs9268861 | 6 | 1.73395E-10 | HLA-DRA |
| rs9268862 | 6 | 1.69701E-15 | HLA-DRA |
| rs9268878 | 6 | 3.02036E-13 | HLA-DRA |
| rs9269186 | 6 | 1.26822E-14 | HLA-DRA |
| rs9270986 | 6 | 1.8543E-08 | HLA-DRB5 |
| rs9271208 | 6 | 9.9083E-08 | HLA-DQA1 |
| rs9271850 | 6 | 1.97E-20 | HLA-DQA1 |
| rs9272219 | 6 | 3.39016E-11 | HLA-DQA1 |
| rs9272723 | 6 | 1.07229E-08 | HLA-DQA1 |
| rs9275418 | 6 | 7.18E-21 | HLA-DQB1 |
| rs9275523 | 6 | 1.13E-28 | HLA-DQB1 |
| rs9275572 | 6 | 1.68E-35 | HLA-DQB1 |
| rs9276435 | 6 | 8.06597E-08 | HLA-DQA2 |
| rs9296009 | 6 | 3.95E-22 | ATF6B |
| rs9348904 | 6 | 1.33218E-08 | HLA-DPA1 |
| rs9368716 | 6 | 4.87E-20 | C6orf10 |
| rs9469240 | 6 | 7.18914E-08 | HLA-DQA2 |
| SNP_A-4251558 (rs4868390) | 5 | 1.80E-24 | SLIT3 |
| SNP_A-4262144 (rs80015580) | 2 | 2.50E-21 | ACOXL |
| SNP_A-4301206 (rs41473844) | 10 | 2.59E-19 | KCNMA1 |
